# Nirsevimab effectiveness against RSV hospitalizations in children 8-23 months at risk of severe RSV

**DOI:** 10.64898/2026.11.13.26368017

**Authors:** Regina M. Simeone, Margaret M. Newhams, Amanda B. Payne, Amber O. Orzel-Lockwood, Jemima Calixte, Natasha B. Halasa, Satoshi Kamidani, Hillary Crandall, Katherine Irby, Aline B. Maddux, Danielle M. Zerr, Judith A. Guzman-Cottrill, Jennifer E. Schuster, Kathleen Chiotos, Michele Kong, Mary Allen Staat, Janet R. Hume, Melissa S. Stockwell, Melissa A. Cameron, Alexander H. Hogan, Jigar C. Chauhan, Austin Biggs, Emily R. Levy, Samina S. Bhumbra, Tracie C. Walker, Neelima K. Marupudi, Mary K. Prahl, Kelly N. Michelson, Lora M. Martin, Tamara T. Bradford, Adrienne G. Randolph, Angela P. Campbell, the Overcoming RSV Network investigators

**Affiliations:** Coronavirus and Other Respiratory Viruses Division, National Center for Immunization and Respiratory Diseases, U.S. Centers for Disease Control and Prevention, Atlanta, Georgia; Department of Anesthesiology, Critical Care, and Pain Medicine, Boston Children’s Hospital, Boston, Massachusetts; Division of Pediatric Infectious Diseases, Department of Pediatrics, Vanderbilt University Medical Center, Nashville, Tennessee; The Center for Childhood Infections and Vaccines of Children’s Healthcare of Atlanta and the Department of Pediatrics, Emory University School of Medicine, Atlanta, Georgia; Division of Pediatric Critical Care, Department of Pediatrics, University of Utah and Primary Children’s Hospital, Salt Lake City, Utah; Section of Pediatric Critical Care, Department of Pediatrics, Arkansas Children’s Hospital, Little Rock, Arkansas; Department of Pediatrics, Section of Critical Care Medicine, University of Colorado School of Medicine and Children’s Hospital Colorado, Aurora, Colorado; Division of Infectious Diseases, Department of Pediatrics, University of Washington, Seattle, Washington; Seattle Children’s Research Institute, Seattle, Washington; Department of Pediatrics, Oregon Health and Sciences University, Portland, Oregon; Division of Pediatric Infectious Diseases, Department of Pediatrics, Children’s Mercy; University of Missouri Kansas City, Kansas City, Missouri; Division of Critical Care Medicine, Department of Anesthesiology and Critical Care, Children’s Hospital of Philadelphia, Philadelphia, Pennsylvania; Division of Pediatric Critical Care Medicine, Department of Pediatrics, University of Alabama at Birmingham, Birmingham, Alabama; Department of Pediatrics, Division of Infectious Diseases, Cincinnati Children’s Hospital Medical Center, University of Cincinnati College of Medicine, Cincinnati, Ohio; Division of Pediatric Critical Care, University of Minnesota Masonic Children’s Hospital, Minneapolis, Minnesota; Division of Child and Adolescent Health, Columbia University Vagelos College of Physicians and Surgeons and Department of Population and Family Health, Columbia University Mailman School of Public Health, New York, New York; Division of Pediatric Hospital Medicine, UC San Diego–Rady Children’s Hospital, San Diego, California; Division of Hospital Medicine, Connecticut Children’s Medical Center, Hartford, Connecticut; Division of Critical Care, Nemours Children’s Hospital, Wilmington, Delaware; Department of Pediatric Critical Care, Medical University of South Carolina and Shawn Jenkins Children’s Hospital, Charleston, South Carolina; Divisions of Pediatric Infectious Diseases and Pediatric Critical Care Medicine, Department of Pediatric and Adolescent Medicine, Mayo Clinic, Rochester, Minnesota; Ryan White Center for Pediatric Infectious Disease and Global Health, Department of Pediatrics, Indiana University School of Medicine, Indianapolis, Indiana; Department of Pediatrics, University of North Carolina at Chapel Hill Children’s Hospital, Chapel Hill, North Carolina; Department of Child Health, University of Arizona, Banner Children’s Hospital at Desert, Division of Pediatric Critical Care, Mesa, Arizona; Division of Pediatric Infectious Diseases, Department of Pediatrics, University of California, San Francisco, California; Division of Pediatric Critical Care Medicine, Ann and Robert H. Lurie Children’s Hospital of Chicago, Chicago, Illinois; Department of Pediatrics, University of Mississippi Medical Center, Jackson, Mississippi; Division of Cardiology, Department of Pediatrics, Louisiana State University Health Sciences Center, Louisiana; Children’s Hospital of New Orleans, New Orleans, Louisiana; Departments of Anesthesia and Pediatrics, Harvard Medical School, Boston, Massachusetts

## Abstract

**Importance:** Respiratory syncytial virus (RSV) is a leading cause of hospitalization in high-risk young children during their second RSV season. Nirsevimab, a long-acting monoclonal antibody against RSV, is recommended for some children who are at increased risk for severe RSV and entering their second RSV season to prevent severe RSV disease. Because of limited eligibility criteria for second season nirsevimab, estimating its effectiveness is challenging.

**Objective:** We evaluated nirsevimab effectiveness against RSV-associated hospitalizations among young children eligible to receive nirsevimab during their second RSV season.

**Design:** This retrospective case control study with a test-negative design was conducted during the 2024–2025 and 2025–2026 RSV seasons.

**Setting:** Enrollment was in 26 hospitals across 24 U.S. states.

**Participants:** Children eligible for nirsevimab during their second RSV season (as recommended in August 2023) were included. Nirsevimab-eligible children 8−23 months old hospitalized with respiratory symptoms were enrolled. Case-patients tested positive for RSV; control-patients tested negative for RSV.

**Exposures:** Receipt of nirsevimab before or during the child’s second RSV season and at least 7 days before symptom onset.

**Outcomes and measures:** RSV-associated hospitalization. We estimated nirsevimab effectiveness using multivariable logistic regression adjusted for age, month and year of hospitalization, census region, and sex comparing the odds of second season nirsevimab receipt among case- and control-patients.

**Results:** One hundred six case-patients and 125 control-patients were analyzed. Distributions of age, sex, race and ethnicity, and region were similar in case- and control-patients. Most case- and control-patients were eligible for nirsevimab because of chronic lung disease of prematurity (84.0% and 69.6%, respectively). Overall, 21 (19.8%) of case-patients and 53 (42.4%) of control-patients received nirsevimab. Adjusted nirsevimab effectiveness against RSV-associated hospitalizations was 78.1% (95% CI, 55.2%-89.3%); the median interval from nirsevimab receipt to symptom onset was 66.5 days (IQR = 30-96 days). Effectiveness was estimated to be higher if nirsevimab was received 8-59 days before symptom onset (88.5%, 95% CI, 63.4%-96.4%).

**Conclusions:** This analysis provides real-world estimates of substantial nirsevimab effectiveness against RSV-associated hospitalizations for high-risk U.S. children in their second RSV season These data support recommendations for nirsevimab as a preventive strategy in this population.

**Key Points:**

- **Question**: What is the effectiveness of nirsevimab against RSV-associated hospitalizations among children 8-23 months old who are at risk for severe disease and in their second RSV season?
- **Findings:** In this case-control study of hospitalizations for acute respiratory illness among high-risk children in their second RSV season, receipt of nirsevimab was associated with an estimated 78% reduction in risk of RSV-associated hospitalizations.
- **Meaning**: Nirsevimab provided substantial protection against RSV-associated hospitalizations in children at high risk of severe RSV during their second RSV season.

## Introduction

Respiratory syncytial virus (RSV) is a leading cause of lower respiratory tract infections (LRTI) and a significant contributor to global hospitalizations for respiratory illness in infants and children.^1,2^ In 2023, two RSV preventive products were approved for use in the United States: a maternal vaccine to be administered during weeks 32 through 36 weeks of gestation,^3^ and nirsevimab, a long-acting monoclonal antibody administered to infants.^4^ Nirsevimab was also approved for certain children up to 19 months of age entering their second RSV season who remain vulnerable to severe RSV disease.^4,5^

As of August 2026, children considered at increased risk of severe disease and recommended to receive nirsevimab in the second season in the U.S. included children of American Indian or Alaskan Native (AI/AN) descent and children with chronic lung disease of prematurity (CLD) requiring medical support in the six months before the RSV season, severe immunocompromise, or cystic fibrosis with manifestations of severe lung disease or weight-for-length <10^th^ percentile (Table 1).^4^ The second season eligibility criteria were chosen based on previously identified conditions that were eligibility criteria to receive palivizumab, a short-acting monoclonal antibody that has since been discontinued in the U.S., and additional evidence indicating severe outcomes among children of AI/AN descent.^4,6,7^

**Table 1.** Factors making a child aged 8-19 months entering their second respiratory syncytial virus season eligible to receive nirsevimab in the United States, as of August 2026^a^.

| **Factor** | **Overcoming RSV condition in the case report form** |
|---|---|
| Chronic lung disease of prematurity requiring ongoing medical support in the 6 months before hospitalization | Bronchopulmonary dysplasia, with receipt of medical support (chronic corticosteroid therapy, diuretic therapy, or supplemental oxygen) in the past 6 months |
| Cystic fibrosis | Cystic fibrosis, with history of hospitalization in the first year of life for a CF pulmonary exacerbation OR persistent abnormalities on chest x-ray or chest CT that persist even when stable and not having pulmonary exacerbations OR weight-for-length <10<sup>th</sup> percentile |
| Severe immunocompromise/<br>immunodeficiency | Any immunosuppressive or oncologic disorder |
| American Indian/Alaska Native or other indigenous population | Race/ethnicity specified as American Indian or Alaska Native |

a. Jones JM, Fleming-Dutra KE, Prill MM, et al. Use of Nirsevimab for the Prevention of Respiratory Syncytial Virus Disease Among Infants and Young Children: Recommendations of the Advisory Committee on Immunization Practices - United States, 2023. MMWR Morb Mortal Wkly Rep. Aug 25 2023;72(34):920-925. doi:10.15585/mmwr.mm7234a4.

Nirsevimab effectiveness against severe RSV-related outcomes has been demonstrated in infants.^1,8,9^ The population of children eligible to receive nirsevimab in their second season was small during the 2024–2025 and 2025–2026 RSV seasons, with eligible criteria impacting approximately 2%-4% of U.S. children^10,11^ As a result, there are few data assessing the effectiveness of nirsevimab in preventing severe RSV disease among high-risk U.S. children in their second RSV season. Nirsevimab has been reported to provide protection against RSV-associated hospitalization among AI/AN children in their second season in Alaska and the Southwest U.S. during 2023–2024,^12,13^ but additional U.S.-based data regarding the effectiveness of nirsevimab in other high-risk populations are limited. Further, clinical efficacy of nirsevimab in a child’s second season was not established during pre-licensure trials, and the second season nirsevimab recommendation relied on serologic data demonstrating non-inferior immunogenicity compared to palivizumab.^6,14^ Estimates of real-world effectiveness of nirsevimab in the second RSV season against severe RSV disease are needed.

Using data from the Overcoming RSV network, we our objective was to estimate the effectiveness of second season nirsevimab immunization against RSV-associated hospitalizations among children aged 8-23 months experiencing their second RSV season at U.S. pediatric hospitals.

## Methods

### Population and enrollment

The Overcoming RSV network, an extension of the Overcoming COVID-19 network, conducts RSV surveillance in 26 pediatric hospitals in 24 states.^8,15^ we used a case-control study with a test-negative design to assess nirsevimab effectiveness among children with conditions that conferred high risk for severe RSV and were 8-23 months of age in their second RSV season. This analysis was limited to children entering their second RSV season (8-19 months of age on October 1^st^ of the 2024–2025 or 2025–2026 RSV seasons) and <24 months at hospitalization during the RSV season. Patients had to be eligible to receive nirsevimab in their second season to be enrolled (Table 1). Hospitalizations were limited to those occurring during RSV season, from November 1, 2024-April 15, 2025, or September 15, 2025-April 15, 2026.

RSV test-positive case-patients were hospitalized for laboratory-confirmed RSV-associated symptoms or signs. RSV test-negative control-patients were hospitalized with RSV-like respiratory symptoms or signs (i.e., cough, shortness of breath, wheezing, retractions, apnea, use of new or elevated oxygen support, or new pulmonary findings on chest imaging consistent with pneumonia) but tested negative for RSV ≤1 day before or during hospitalization. Case- and control-patients were matched by site of enrollment, age category (8-15 months, 16-23 months), and hospital admission date (+/− 2 months). Up to two control-patients were identified per case-patient. Case and control symptom onset for the current respiratory illness had to have begun within 14 days of hospitalization. This activity was reviewed by CDC, deemed not research, and was conducted consistent with applicable federal law and CDC policy (See e.g., 45 C.F.R. part 46, 21 C.F.R. part 56; 42 U.S.C. §241(d); 5 U.S.C. §552a; 44 U.S.C. §3501 et seq.). The investigation followed the Strengthening the Reporting of Observational Studies in Epidemiology (STROBE) reporting guidelines.^16^

### Data collection

Demographic characteristics and clinical illness information were obtained through caregiver interview and medical record abstraction. Information about second season nirsevimab-eligible and other preexisting health conditions was obtained through medical record abstraction. Information about gestational age at delivery was collected for infants <12 months of age at admission. Nirsevimab receipt was defined as having received one dose ≥7 days before symptom onset and after the end of the child’s first RSV season. Receipt was considered verified if documentation of a nirsevimab dose was confirmed in the child’s medical record or the state’s immunization information system, or if plausible receipt of nirsevimab was indicated in the caregiver interview and an exact or approximate date of receipt was provided.

Patients were not eligible for inclusion if nirsevimab receipt occurred within 7 days of symptom onset or during hospitalization, if symptom onset began more than 14 days before hospital admission, if nirsevimab receipt was not verified or did not have plausible caregiver-report, if the incorrect nirsevimab dose was received, if sites could not access the child’s medical record or immunization information system record, if the patient did not have an RSV test result, or if they did not have a nirsevimab eligible condition.

### Analysis

We compared demographic and clinical characteristics between case- and control-patients and patients who did and did not receive second season nirsevimab using chi-squared, Fisher’s exact, or Kruskal-Wallis tests. We calculated effectiveness of second season nirsevimab against RSV-associated hospitalization using logistic regression, comparing the odds of nirsevimab receipt among case- and control-patients; effectiveness was calculated as 100%×(1-adjusted odds ratio). Models were adjusted for age (in months), census region (Northeast, South, Midwest, and West), sex, month/year of hospitalization, and presence of a medical condition other than one conferring nirsevimab eligibility; site was included as a random effect.

Sites attempted to match control-patients to case-patients on age group and date of admission (+/− two months) in a two-to-one ratio. However, because matching was unsuccessful for some cases, an unmatched complete case analysis was conducted as the primary model to assess nirsevimab effectiveness. For all patients, we also assessed effectiveness of nirsevimab received in the 8-59 days before symptom onset, or ≥60 days before symptom onset. A matched analysis, restricting to informative matched groups of case- and control-patients was conducted as a sensitivity analysis. As an additional set of sensitivity analyses, we excluded case- and control-patients who tested positive for influenza or SARS-CoV-2, and evaluated effectiveness overall, by time since symptom onset, and in a matched analysis. For all statistical analyses, p-values <0.05 were considered statistically significant.

## Results

We enrolled 244 children 8-23 months of age with a hospitalization date between November 1, 2024-April 15, 2025, or September 15, 2025-April 15, 2026. After exclusion of 13 children, a total of 106 case-patients 8-23 months of age hospitalized for RSV during their second RSV season and 125 control-patients who met U.S. eligibility criteria for nirsevimab receipt during their second RSV season were included (Supplemental Figure 1). Median age for case-patients was 15.5 months (interquartile range [IQR]: 13.6-18.7 months); for control-patients, median age was 15.9 months (IQR: 13.4, 18.5 months) (Table 2). There were no statistically significant differences in the distribution of race and ethnicity, region, and social vulnerability index between case- and control-patients.

**Table 2.** Characteristics of case-patients with respiratory syncytial virus (RSV) and control-patients testing negative for RSV, and among those who did and did not receive second season nirsevimab, among nirsevimab eligible children 8 through 23 months of age during their second RSV season, November 1, 2024-April 15, 2025 and September 15, 2025-April 15, 2026, Overcoming RSV Network

| Characteristic | Total<br>No. (%) | RSV+ case-patients<br>No. (%) | RSV- control-patients<br>No. (%) | P-value<sup>a</sup> | Received nirsevimab<br>No. (%) | Did not receive nirsevimab<br>No. (%) | P-value<sup>a</sup> |
|---|---|---|---|---|---|---|---|
| **Total** | **231** | **106** | **125** |  | **74** | **157** |  |
| **RSV season<sup>b</sup>** |  |  |  |  |  |  |  |
| 2024-25 season | 128 (55.4) | 58 (54.7) | 70 (56.0) | 0.85 | 41 (55.4) | 87 (55.4) | 0.999 |
| 2025-26 season | 103 (44.6) | 48 (45.3) | 55 (44.0) |  | 33 (44.6) | 70 (44.6) |  |
| **Age - median (IQR)** | 15.8 (13.4, 18.5) | 15.5 (13.6, 18.7) | 15.9 (13.4, 18.5) | 0.92 | 15.3 (12.7, 18.4) | 15.9 (14.0, 18.7) | 0.92 |
| **Sex** |  |  |  |  |  |  |  |
| Male | 125 (54.1) | 62 (58.5) | 63 (50.4) | 0.22 | 40 (54.1) | 85 (54.1) | 0.99 |
| Female | 106 (45.9) | 44 (41.5) | 62 (49.6) |  | 34 (45.9) | 72 (45.9) |  |
| **Race and ethnicity<sup>c</sup>** |  |  |  |  |  |  |  |
| Non-Hispanic White | 74 (32.0) | 31 (29.2) | 43 (34.4) | 0.68 | 26 (35.1) | 48 (30.6) | 0.37 |
| Non-Hispanic Black | 50 (21.6) | 25 (23.6) | 25 (20.0) |  | 22 (29.7) | 28 (17.8) |  |
| Non-Hispanic Asian | 9 (3.9) | 4 (3.8) | 5 (4.0) |  | 2 (2.7) | 7 (4.5) |  |
| Hispanic | 45 (19.5) | 25 (23.6) | 20 (16.0) |  | 11 (14.9) | 34 (21.7) |  |
| Multiple or other | 9 (3.9) | 3 (2.8) | 6 (4.8) |  | 2 (2.7) | 7 (4.5) |  |
| American Indian/Alaska Native | 29 (12.6) | 10 (9.4) | 19 (15.2) |  | 5 (6.8) | 24 (15.3) |  |
| Unknown | 15 (6.5) | 8 (7.5) | 7 (5.6) |  | 6 (8.1) | 9 (5.7) |  |
| **Region<sup>d</sup>** |  |  |  |  |  |  |  |
| Northeast | 20 (8.7) | 10 (9.4) | 10 (8.0) | 0.44 | 6 (8.1) | 14 (8.9) | 0.42 |
| Midwest | 25 (10.8) | 15 (14.2) | 10 (8.0) |  | 8 (10.8) | 17 (10.8) |  |
| South | 111 (48.1) | 47 (44.3) | 64 (51.2) |  | 41 (55.4) | 70 (44.6) |  |
| West | 75 (32.5) | 34 (32.1) | 41 (32.8) |  | 19 (25.7) | 56 (35.7) |  |
| **Social vulnerability index – median (interquartile range)** | 0.5 (0.3, 0.7) | 0.5 (0.3, 0.7) | 0.5 (0.4, 0.7) | 0.59 | 0.5 (0.4, 0.7) | 0.5 (0.3, 0.7) | 0.32 |
| **Gestational age (n=28)<sup>e</sup>** |  |  |  |  |  |  |  |
| <37 weeks gestation | 19/28 (67.9) | 9/12 (75.0) | 10/16 (62.5) | 0.69 | 9/12 (75.0) | 10/16 (62.5) | 0.69 |
| ≥ 37 weeks gestation | 9/28 (32.1) | 3/12 (25.0) | 6/16 (37.5) |  | 3/12 (25.0) | 6/16 (37.5) |  |
| **Nirsevimab eligibility<sup>f</sup>** |  |  |  |  |  |  |  |
| Chronic lung disease of prematurity | 176 (76.2) | 89 (84.0) | 87 (69.6) | 0.01 | 67 (90.5) | 109 (69.4) | <.001 |
| Severe immunocompromise | 29 (12.6) | 7 (6.6) | 22 (17.6) | 0.01 | 3 (4.1) | 26 (16.6) | 0.01 |
| American Indian/Alaska Native | 29 (12.6) | 10 (9.4) | 19 (15.2) | 0.19 | 5 (6.8) | 24 (15.3) | 0.07 |
| Cystic fibrosis | 5 (2.2) | 3 (2.8) | 2 (1.6) | 0.66 | 1 (1.4) | 4 (2.5) | 0.67 |
| **Preexisting health conditions** |  |  |  |  |  |  |  |
| Any underlying condition<sup>g</sup> | 141 (61.0) | 67 (63.2) | 74 (59.2) | 0.53 | 58 (78.4) | 83 (52.9) | <.001 |
| Neuromuscular or neurologic condition<sup>h</sup> | 46 (19.9) | 15 (14.2) | 31 (24.8) | 0.04 | 17 (23.0) | 29 (18.5) | 0.42 |
| Respiratory condition<sup>i</sup> | 87 (37.7) | 36 (34.0) | 51 (40.8) | 0.29 | 40 (54.1) | 47 (29.9) | <.001 |
| Cardiac condition<sup>j</sup> | 75 (32.5) | 31 (29.2) | 44 (35.2) | 0.34 | 36 (48.6) | 39 (24.8) | <.001 |
| Metabolic, genetic, or endocrine<sup>k</sup> | 39 (16.9) | 15 (14.2) | 24 (19.2) | 0.31 | 15 (20.3) | 24 (15.3) | 0.35 |
| **Nirsevimab** |  |  |  |  |  |  |  |
| Nirsevimab in second season | 74 (32.0) | 21 (19.8) | 53 (42.4) | 0.12 | 74 (100.0) | N/A | N/A |
| Days from nirsevimab receipt to symptom onset- median (IQR) | 66.5 (30.0, 96.0) | 75.0 (64.0, 107.0) | 64.0 (29.0, 94.0) | <.001 | 66.5 (30.0, 96.0) | N/A | N/A |
| First season nirsevimab | 117 (50.6) | 55 (51.9) | 62 (49.6) | 0.73 | 41 (55.4) | 76 (48.4) | 0.32 |
| **Highest level of respiratory support** |  |  |  |  |  |  |  |
| Low flow or no respiratory support | 102 (45.9) | 40 (38.8) | 62 (52.1) | 0.01 | 34 (50.0) | 68 (44.2) | 0.01 |
| High flow nasal cannula | 51 (23.0) | 25 (24.3) | 26 (21.8) |  | 11 (16.2) | 40 (26.0) |  |
| Noninvasive ventilation<sup>l</sup> | 42 (18.9) | 29 (28.2) | 13 (10.9) |  | 8 (11.8) | 34 (22.1) |  |
| Invasive mechanical ventilation | 27 (12.2) | 9 (8.7) | 18 (15.1) |  | 15 (22.1) | 12 (7.8) |  |
| **Clinical course** |  |  |  |  |  |  |  |
| ICU admission | 105 (45.5) | 49 (46.2) | 56 (44.8) | 0.83 | 33 (44.6) | 72 (45.9) | 0.86 |
| ECMO | 2 (0.9) | 1 (0.9) | 1 (0.8) | 1.00 | 1 (1.4) | 1 (0.6) | 0.54 |
| Death during hospitalization | 1 (0.4) | 0 | 1 (0.8) | 1.00 | 0 | 1 (0.6) | 1.00 |
| Hospital length of stay | 5.0 (4.0, 8.0) | 6.5 (5.0, 11.0) | 4.0 (3.0, 7.0) | <.001 | 5.0 (3.0, 9.0) | 5.0 (4.0, 8.0) | 0.92 |

Abbreviations: ECMO, extracorporeal membrane oxygenation; ICU, intensive care unit; IQR, interquartile range; RSV, respiratory syncytial virus
a. Chi-squared p-value to assess differences in proportions across groups; comparisons with expected cell counts <5 in >20% of cells used Fisher’s exact test; comparisons of continuous variables used Kruskal-Wallis tests
b. 2024-25 season among children admitted between November 1, 2024-April 15, 2025 and September 15, 2025-April 15, 2026
c. Statistical testing did not include American Indian/Alaska Native children
d. Northeast: Connecticut Children's Medical Center, Boston Children’s Hospital, Columbia University, Children’s Hospital of Philadelphia; Midwest: Ann & Robert H. Lurie Children's Hospital of Chicago, Riley Hospital for Children, Mayo Clinic Children’s, University of Minnesota Masonic Children’s Hospital, Children's Mercy Kansas City, Cincinnati Children's Hospital Medical Center; South: Children's of Alabama, Arkansas Children's Hospital, Nemours Children’s Hospital, Children’s Healthcare of Atlanta, Manning Family Children's, UNC Children’s Hospital, Medical University of South Carolina, Monroe Carell Jr. Children's Hospital at Vanderbilt; West: Arizona Mesa Children’s Hospital, UCSF Benioff Children's Hospital, Children's Hospital Colorado, Doernbecher Children’s Hospital, Primary Children’s Hospital, Rady Children’s Hospital, Seattle Children’s Hospital
e. Gestational age at birth was assessed among children <12 months of age at hospitalization
f. Eligibility criteria as of August 2026
g. Summary group of any of respiratory, cardiac, neurological/neuromuscular, and metabolic/genetic/endocrine conditions
h. Neuromuscular or neurologic conditions included muscular dystrophy, static encephalopathy, spastic quadriplegia, seizure disorders, neuromuscular weakness, and hydrocephalus
i. Respiratory conditions included asthma, requiring assistance with clearance of secretions, and upper airway disorders
j. Cardiac conditions included congenital heart disease, history of cardiac repair, pulmonary hypertension, and congestive heart failure
k. Metabolic, genetic, or endocrine conditions included trisomy 21, other chromosomal disorders, mitochondrial disorders, diabetes, and other metabolic disorders
l. Noninvasive ventilation defined as receipt of bilevel positive airway pressure or continuous positive airway pressure

The majority (n=176, 76.2%) of all case- and control-patients eligible for second season nirsevimab qualified due to CLD (89 case- and 87 control-patients) (Table 2), followed by having severe immunocompromise (n=29; 7 case- and 22 control-patients), being AI/AN (n=29; 10 case- and 19 control-patients), and cystic fibrosis (n=5; 3 case- and 2 control-patients). Two AI/AN patients also had a severely immunocompromising condition, four AI/AN patients also had CLD, and two patients with CLD also had severe immunocompromise. By case-control status, 84.0% of case-patients, and 69.6% of control-patients had CLD requiring medical support in the six months before the RSV season (p=0.01) (Table 2). A lower proportion of case-patients had severe immunocompromise (6.6%) than control-patients (17.6%) (p=0.01). Being eligible due to cystic fibrosis or being AI/AN had similar proportions between case- and control-patients. Outside of conditions making children eligible for nirsevimab, a greater proportion of control-patients had neuromuscular or neurologic conditions (24.8%) than case-patients (14.2%) (p=0.04). There were no other statistically significant differences in underlying conditions by case- and control-patient status.

A significantly higher proportion of control-patients received second season nirsevimab (42.4%) than case-patients (19.8%, p<0.001) (Table 2). Median length of time from nirsevimab receipt to symptom onset was longer among case-patients than control-patients (case-patients: 75.0 days; control-patients: 64.0 days) (p<0.001). The maximum number of days between nirsevimab receipt and symptom onset for case- and control-patients was 178 days; 12 patients (5 case- and 7 control-patients) received nirsevimab ≥120 days before symptom onset. Respectively, 51.9% and 49.6% of case- and control-patients had received a first season dose of nirsevimab.

There were no statistically significant differences in second season nirsevimab receipt by season, age, sex, race and ethnicity, region, or social vulnerability index (Table 2). Patients who received second season nirsevimab were more likely to have CLD (90.5% vs. 69.4%, p<0.001) and less likely to have severe immunocompromise (4.1% vs. 16.6%, p=0.01). Patients who received second season nirsevimab were also more likely to have an additional, non-nirsevimab eligible preexisting health condition (78.4% vs. 52.9%, p<0.001); respiratory and cardiac conditions were more common.

Among case-patients, those who received second season nirsevimab were more likely to also have a non-nirsevimab eligible underlying condition (p=0.02), in addition to the condition that made them eligible, than those who had not received nirsevimab (Supplemental Table 1). These additional conditions most commonly included respiratory (52.4%) and cardiac (42.9%) conditions. A higher proportion of a subset of RSV-positive case-patients who received nirsevimab and were clinically tested for additional pathogens also had co-infections with other respiratory viruses (28.6% vs. 20.0%) or bacterial pathogens (33.3% vs. 10.7%) compared with case-patients who did not receive nirsevimab, but these differences were not statistically significant.

In the main analysis using unmatched data, overall adjusted estimated nirsevimab effectiveness against second-season RSV hospitalization was 78.1% (95% CI, 55.2%-89.3%) (Figure). Point estimates of adjusted nirsevimab effectiveness were higher among patients that received nirsevimab 8-59 days prior to hospitalization (88.5% [95% CI, 63.4%-96.4%]) than among patients that received nirsevimab 60-178 days prior to hospitalization (69.8% [32.1%-86.5%]), although confidence intervals overlapped. In the sensitivity analysis including only matched case- and control-patients, estimated adjusted nirsevimab effectiveness was 76.1% (95% CI, 41.8%-90.2%). In the sensitivity analysis excluding patients with an influenza or SARS-CoV-2 infection (three case- and 17 control-patients), the distribution of patient characteristics by case- and control-patient status and nirsevimab receipt were similar (Supplemental Table 2). Nirsevimab effectiveness estimates were also similar to the main analysis, with overall effectiveness of 80.7% (95% CI, 59.2%-96.6%) (Supplemental Figure 2), although patients receiving nirsevimab in the 60-178 days prior to hospitalization had slightly increased effectiveness (76.4% [45.7%-80.7%]).

## Discussion

In this multi-site analysis of children in their second RSV season with conditions making them eligible to receive nirsevimab during the 2024–2025 and 2025–2026 RSV seasons,^4^ nirsevimab was estimated to be 78.1% effective at preventing RSV-associated hospitalizations. Median time from nirsevimab receipt to symptom onset among case-patients was 75 days, and nirsevimab received in the 59 days before symptom onset was estimated to be 88.5% effective at preventing RSV-associated hospitalizations. Patients who received second season nirsevimab 60-178 days after receipt had 69.8% lower risk of being hospitalized with RSV than those who did not receive nirsevimab.

The conditions conferring second season nirsevimab eligibility in these two seasons in the U.S. are rare, making it difficult to assess nirsevimab effectiveness because of small sample size.^10^ The second-season nirsevimab effectiveness observed in the current analysis of the 2024-2025 and 2025-2026 seasons is similar to that observed for infants in their first RSV season in other populations. For example, in the New Vaccine Surveillance Network, nirsevimab was 93% effective (95% CI: 82%-97%) at preventing hospitalization.^1^ Likewise, nirsevimab was 80% effective against first season pediatric intensive care unit admission in the Overcoming RSV network (95% CI: 70%–86%).^8^ First season nirsevimab is recommended for all infants <8 months entering their first RSV season who are not already protected by maternal vaccination.^1,4,8^

As for data in the second season, a U.S. study of second season nirsevimab effectiveness among AI/AN children during 2023-2024 reported that nirsevimab was 88% (95% CI: 22%-98%) effective against RSV hospitalization.^12^ In a different assessment of second season nirsevimab effectiveness in Alaska’s Yukon-Kuskokwim Delta region in 2023-2024, nirsevimab was 88% effective against medically attended RSV illness in the second season (95% CI: 48%-97%).^13^ These higher effectiveness estimates are likely due to differences in the underlying population, with fewer children having high-risk health conditions when restricted to those who are AI/AN compared to the present analysis. A 2026 Spanish study observed a lower second season nirsevimab effectiveness estimate of 42% (95% CI: −25%-73%),^17^ although the estimate was underpowered and lacked precision. This lower point estimate may be due to broader second season nirsevimab eligibility criteria or a lower threshold for hospitalization used in Spain, potentially attenuating effectiveness. These findings from our large multisite network estimating effectiveness of second season nirsevimab in young children with high-risk conditions fill a critical data gap for this vulnerable population.

All infants are recommended to be protected by either maternal RSV vaccination or receipt of monoclonal RSV antibody to prevent severe RSV during their first RSV season. Residual immunity from products used in the first RSV season is not well established for the second season and is unlikely to be meaningful.^18^ At least four studies have examined whether nirsevimab received in the first season is protective in the second season, with two studies reporting reduction in risk of RSV-related LRTI hospitalizations in the second season.^19–22^ These studies did not include all high-risk patients. In our analysis, approximately half of case- and control-patients had received nirsevimab in the first season, so this was unlikely to be an important factor influencing second season protection.

We observed in our network that RSV-positive children who received nirsevimab in their second season were more likely to have additional underlying health conditions than those RSV-positive children who did not receive second season nirsevimab. These additional comorbidities may have contributed to the need for hospitalization in some children. In addition, although patients were not tested systematically, coinfections with additional respiratory viral and bacterial pathogens were present among more than a quarter (28.6%) of the subset of nirsevimab-recipient case-patients tested.

These estimates among children 8 through 23 months represent effectiveness estimated among those children experiencing RSV circulation during the first three years of U.S. nirsevimab administration. During the 2023-2024 RSV season, nirsevimab coverage among infants <8 months was 18.5,%^23^ and increased to 44.7% in the 2024-2025 season.^24–26^ Changes in the underlying epidemiology and burden of RSV may occur due to increased coverage of maternal RSV vaccination and nirsevimab use during the first RSV season. Understanding how second season nirsevimab effectiveness may change when large proportions of the population are protected in the first season by RSV preventive products before entering their second season, and when nirsevimab coverage is higher among those in their second season, will be important.

### Strengths and limitations

This study has several strengths. This is one of the first U.S. studies able to estimate second season nirsevimab effectiveness in this select population of children with conditions that place them at high-risk of severe RSV. It used data from a large, heterogeneous population contributed by 26 U.S. hospitals, with enrollment using detailed medical record review and abstraction to identify nirsevimab-eligible conditions, and verification of nirsevimab receipt in order to reduce misclassification. Further, using a strict clinical definition based on respiratory symptoms and signs for case- and control-patients, the test-negative design allowed control-patient selection from the same source population as the case-patients, reducing issues of confounding and selection bias.^27,28^ Because the conditions making a child eligible for nirsevimab receipt are rare in the population,^10^ conducting a randomized clinical trial of second season nirsevimab to evaluate its efficacy would be infeasible due to difficulty recruiting sufficient numbers of eligible patients.^29^

However, this study is subject to several limitations. First, despite having data from two RSV seasons in this surveillance system, there were still relatively small numbers of case- and control-patients eligible to receive nirsevimab during their second RSV season. Those patients enrolled in this analysis were predominantly eligible because they had chronic lung disease of prematurity. It was not possible to assess effectiveness by specific underlying conditions conferring eligibility or to conduct subgroup analyses. Second, clinical RSV testing was used to identify case- and control-patients, but systematic testing for coinfections with respiratory pathogens was not conducted; therefore, we could not assess the impact of other pathogens on these results. Third, because this was an observational test-negative study, it is subject to potential residual confounding; however, the results from the matched analysis were similar to the complete case analysis, providing some reassurance. Fourth, this analysis was restricted to those eligible to receive nirsevimab during their second RSV season as of the time of study enrollment during the 2024–2025 and 2025–2026 RSV seasons,^6,30^ and effectiveness among other children potentially at high risk of severe RSV was not assessed. It is possible that children with other conditions known to increase the risk of severe RSV could benefit from nirsevimab during their second RSV season.^31–35^

## Conclusions

In this case-control study using a test-negative design during the 2024–2025 and 2025–2026 RSV seasons, these data provide real world evidence that receipt of nirsevimab provided protection against RSV-associated hospitalizations for high-risk nirsevimab-eligible children in their second RSV season. These estimates support the ongoing recommendation for use of nirsevimab as a preventive strategy in high-risk young children during their second RSV season.

## Supporting information

Supplemental content

## Data Availability

Data are not publicly available. Deidentified participant data in a limited dataset may be made available upon formal request for a specified purpose for researchers whose proposed use of the data has been approved.

## Overcoming RSV Investigators

**Alabama:** <u>Children’s of Alabama, Birmingham.</u> Michele Kong, MD; Meghan Murdock, RN; Heather Kelley, RN; Candice Colston

**Arizona:** <u>Banner Children’s Hospital, Mesa.</u> Neelima Marupudi, MD

**Arkansas:** <u>Arkansas Children’s Hospital, Little Rock.</u> Katherine Irby, MD; Ronald C. Sanders, MD; Masson Yates, RN; Ashlyn Madding, RN, MSN

**California:** <u>Rady Children’s Hospital, San Diego.</u> Melissa A. Cameron, MD, Michael Henne

**California:** <u>UCSF Benioff Children’s Hospital, San Francisco.</u> Mary Prahl, MD, Nadia Bourdoud, MPH, CRC

**Colorado:** <u>Children’s Hospital Colorado, Aurora.</u> Aline B. Maddux, MD, MSCS, Ariana Valenzuela, CRC, Natasha Baig, MBBS, CRC, Lexi Petruccelli, CRC, Rachel Greer, CRC, Lanae Dailey, RRT, CRC, Heidi Sauceda, EdS, MS, Sara Shankman, CPNP-AC, DNP, Jaime Laurin, MS

**Connecticut:** <u>Connecticut Children’s Medical Center, Hartford.</u> Alexander H. Hogan, MD, MS; Katherine Herbst; Cameron King

**Delaware:** <u>Nemours Children’s Hospital, Wilmington, Delaware; Sidney Kimmel Medical College, Thomas Jefferson University, Philadelphia, PA.</u> Jigar C. Chauhan, MD; Scott L. Weiss, MD; Rebecca Clifford; Jenna Lapira

**Georgia:** <u>Emory University School of Medicine and Children’s Healthcare of Atlanta,</u> <u>Atlanta.</u> Satoshi Kamidani, MD, PhD; Mark D. Gonzalez, PhD; Caroline R. Ciric, BS; Jong-Ha C. Choi, BA; Gabriella Ess, BS; Anna K. Mitchell, BS; Luis W. Salazar, MPH; Janelle J. Spencer-Ramirez, BIS. Humerazehra Momin, BIS.

**Illinois:** <u>Ann & Robert H. Lurie Children’s Hospital of Chicago, Chicago.</u> Kelly N. Michelson, MD, MPH; Bria M. Coates, MD; Sarah Jae Wershil, BA; Alexandra Reyes, BS.

**Indiana:** <u>Riley Hospital for Children, Indianapolis.</u> Samina S. Bhumbra, MD; Mary Stumpf, MS, CCRC.

**Louisiana:** <u>Children’s Hospital of New Orleans, New Orleans.</u> Tamara T. Bradford, MD.

**Massachusetts:** <u>Boston Children’s Hospital, Boston.</u> Adrienne G. Randolph, MD; Margaret M. Newhams, MPH; Amber O. Orzel-Lockwood, MPH; Jemima M. Calixte, MS; Amira Toivonen; Callum Ackerman.

**Minnesota:** <u>University of Minnesota Masonic Children’s Hospital, Minneapolis,</u> Janet R. Hume, MD, PhD; Joanna Ly, CRC; Judith Kyrkos, CRC.

**Minnesota:** <u>Mayo Clinic, Rochester.</u> Emily R. Levy, MD; Chloe A Schluttner; Carly R Crowley

**Mississippi:** <u>Children’s Hospital of Mississippi, Jackson.</u> Lora Martin, MSN; Joseph M. Majure, MD; Lacy Malloch, BS; Maygan Martin, RN

**Missouri:** <u>Children’s Mercy Kansas City, Kansas City.</u> Jennifer E. Schuster, MD; Berenice Marrufo, CCRP

**New York:** <u>Columbia University Irving Medical Center, New York City.</u> Melissa S. Stockwell, MD MPH; Tom Connors, MD; Celibell Y. Vargas; Raul A. Silverio Francisco; Ana Valdez de Romero

**North Carolina:** <u>University of North Carolina at Chapel Hill, Chapel Hill.</u> Stephanie P. Schwartz, MD; Tracie C. Walker, MD; Alison Mathers

**Ohio:** <u>Cincinnati Children’s Hospital, Cincinnati.</u> Mary Allen Staat, MD, MPH; Chelsea C. Rohlfs, BS, MBA; Elizabeth P. Schlaudecker, MD, MPH

**Oregon:** <u>Oregon Health and Science University, Portland.</u> Judith A. Guzman-Cottrill, DO; Chelsea Heisler, CCRP.

**Pennsylvania:** <u>Children’s Hospital of Philadelphia, Philadelphia.</u> Kathleen Chiotos, MD, MSCE; Alanah Mckelvey, BS; Jenny Bush, RN, BSN; Rebecca Douglas, RN, BSN; Myooran Sivarupan, BS; Glory Edioma, BA; Taylor A. Slocumb, BA; Erin McDowell, MA

**South Carolina:** <u>MUSC Children’s Health, Charleston.</u> Austin Biggs, MD; Zach Rusler

**Tennessee:** <u>Monroe Carell Jr. Children’s Hospital at Vanderbilt, Nashville.</u> Natasha B. Halasa, MD, MPH; Laura S. Stewart, PhD; Kristina A. Betters, MD; Annika Conlee; Madeline Spradley; Adriana Blanco Vasquez, Eliana Rebolledo

**Utah:** <u>University of Utah and Primary Children’s Hospital, Salt Lake City.</u> Hillary Crandall, MD, PhD; Kendall Armstrong

**Washington:** <u>Seattle Children’s Hospital, Seattle.</u> Danielle M. Zerr, MD, MPH; Amanda Adler.

## Conflicts of interest

This work was funded by the Centers for Disease Control and Prevention (CDC) under a contract (75D30122C13330) with Boston Children’s Hospital (Randolph, Newhams, Orzel-Lockwood, Calixte). All participating institutions received payment for patient enrollment through a sub-contract with Boston Children’s Hospital and Harvard Medical School.

No conflicts: Campbell, Payne, Simeone

Biggs, Bradford, Chauhan, Chiotos, Guzman-Cottrill, Martin, Marupudi, Walker, Zerr report receipt of payment from CDC to their institution through a subcontract with Boston Children’s Hospital and Harvard Medical School

Crandall: Grants or contracts from NIH/NIGMS to institution

Halasa: Grants or contracts from Sanofi and royalties or licenses from UpToDate

Hogan: NIH/NIAID K award; payment for expert testimony from Mortensen and Milne to institution;

Hume: Grants or contracts from NIH and CDC to institution; membership on the Executive Board, Section on Critical Care of American Academy of Pediatrics (unpaid)

Irby: Grants or contracts from NIH to institution

Kamidani: Grants or contracts from NIH, Pfizer, Moderna, Meissa, Bavarian Nordic, and Sanofi to institution; payment or honoraria from American Academy of Pediatrics

Levy: Grants or contracts from the Pediatric Pandemic Network/HRSA to institution

Michelson: Grants of contracts from PCORI, NOH, NSF; support for attending the AAP NCE meeting; leadership on the AAP Committee on Bioethics and Normal Moment; cofounder of Missing Pieces

Prahl: Grants from NIH and CDC

Randolph: Contract from CDC; Grants or contracts from NIH, NIAID; Royalties or licenses from Up to Date; Participation in the boards of NHLBI Prevent VILI and REMAP CAP; leadership or fiduciary role as an International Sepsis Forum Council Member

Schuster: Grants from NIH, CDC, FDA, State of Missouri to Institution; consulting fees from the Association of Professionals in Infection Control and Epidemiology; Speaking honoraria from Missouri American Academy of Pediatrics; Review panel payment from CDC; Support for attending meetings and/or travel from the Infectious Diseases Society of American and the Pediatric Infectious Diseases Society; Advisory board membership on the Association of American Medical Colleges; Leadership role on the Pediatric Infectious Diseases Society Board.

Staat: Grants or contracts from CDC, NIH, Cepheid, and Merck; Royalties or licenses from Up to Date; Consulting fees from Merck

Stockwell: Grants or contracts from Westat and monoclonal antibodies paid to institution; leadership or fiduciary role as Associate Director of the American Academy of Pediatrics’ Pediatric Research in Office Settings

## Funding/Acknowledgements

This work was funded by CDC through contract # 75D30122C13330 to Boston Children’s Hospital.

## Disclaimer

The findings and conclusions in this report are those of the authors and do not necessarily represent the official position of the Centers for Disease Control and Prevention.

---

**Figure.**
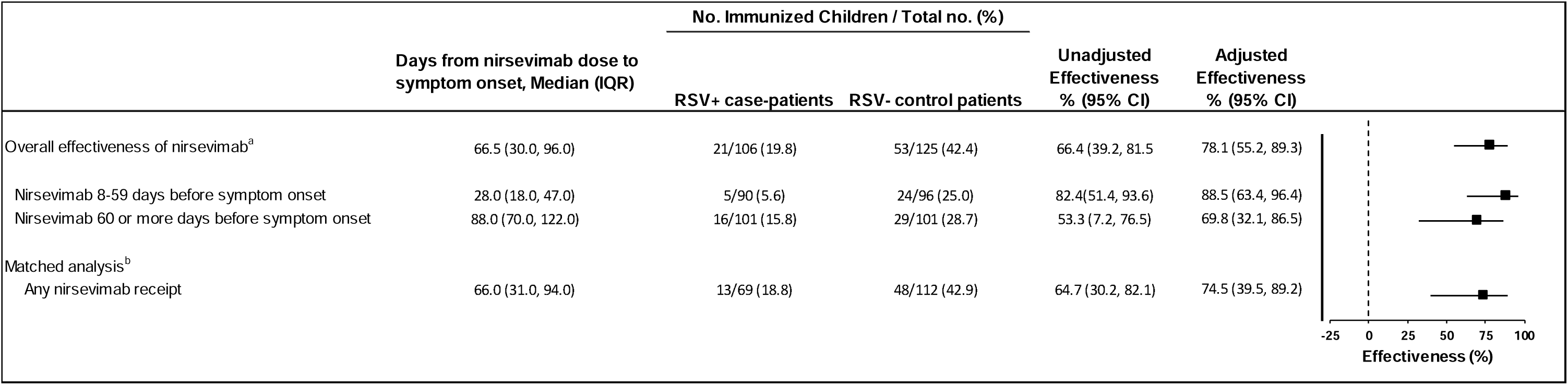
Second season nirsevimab effectiveness against respiratory syncytial virus-associated hospitalizations among nirsevimab-eligible children aged 8-23 months during their second RSV season, November 1, 2024-April 15, 2025 and September 15, 2025-April 15, 2026, Overcoming RSV Network Abbreviations: CI, confidence interval; IQR, interquartile range; RSV, respiratory syncytial virus a. Effectiveness (%) was calculated as (100*(1-adjusted odds ratio), with adjusted odds ratio calculated using unconditional logistic regression and adjusted for age, medical conditions other than nirsevimab, region, month/year of hospitalizations, and sex; controlled for site as a random effect. Data from the Overcoming RSV network are a case control study with a test negative design. b. Effectiveness (%) was calculated as (100*(1-adjusted odds ratio), with adjusted odds ratio calculated using conditional logistic regression and adjusted for age, medical conditions other than nirsevimab, month/year of hospitalization, and sex. Data from the Overcoming RSV network are a case control study with a test negative design.

