## Supplemental content for "Nirsevimab effectiveness against RSV hospitalizations in children 8-23 months at risk of severe RSV"

Supplemental Figure 1. Enrollment and inclusion of case-patients with respiratory syncytial virus (RSV) and control-patients testing negative for RSV, among children 8 through 23 months of age during their second RSV season, November 1, 2024-April 15, 2025 and September 15, 2025-April 15, 2026, Overcoming RSV Network

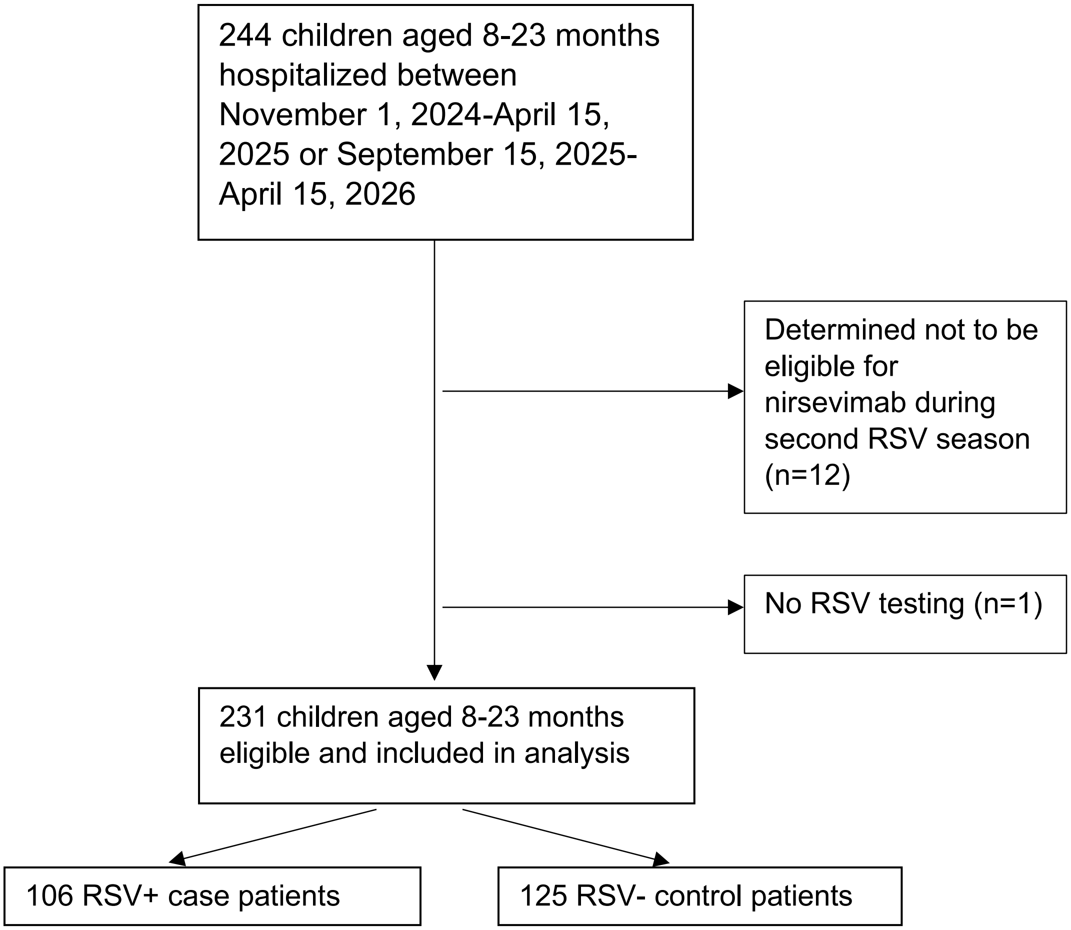

| Supplemental Table 1. Characteristics of case-patients with respiratory syncytial virus (RSV) who did and did not receive nirsevimab during their second RSV season, among nirsevimab-eligible children 8 through 23 months of age during their second RSV season, November 2024-March 2025 and October 2025-April 2026, Overcoming RSV Network | | | |
|---|---|---|---|
| | **RSV+ Case patients** | | |
| **Characteristic** | **Received nirsevimab No. (%)** | **Did not receive nirsevimab No. (%)** | **P-value[a]** |
| **Total** | 21 | 85 | |
| **RSV season[b]** | | | |
| 2024-25 season | 11 (52.4) | 47 (55.3) | 0.81 |
| 2025-26 season | 10 (47.6) | 38 (44.7) | |
| **Age - Median (IQR)** | 1.3 (1.1, 1.5) | 1.3 (1.2, 1.6) | 0.27 |
| **Sex** | | | |
| Male | 15 (71.4) | 47 (55.3) | 0.18 |
| Female | 6 (28.6) | 38 (44.7) | |
| **Gestional age (n=12)[c]** | | | |
| <37 weeks gestation | 4/4 (100.0) | 5/8 (62.5) | 0.49 |
| ≥ 37 weeks gestation | | 3/8 (37.5) | |
| **Nirsevimab eligibility[d]** | | | |
| Chronic lung disease of prematurity | 20 (95.2) | 69 (81.2) | 0.18 |
| Severe immunocompromise | 0 | 7 (8.2) | 0.34 |
| American Indian/Alaska Native | 0 | 10 (11.8) | 0.21 |
| Cystic fibrosis | 1 (4.8) | 2 (2.4) | 0.49 |
| **Preexisting health conditions, not nirsevimab eligible** | | | |
| Any underlying condition[e] | 18 (85.7) | 49 (57.6) | 0.02 |
| Neuromuscular or neurologic condition[f] | 2 (9.5) | 13 (15.3) | 0.73 |
| Respiratory condition[g] | 11 (52.4) | 25 (29.4) | 0.05 |
| Cardiac condition[h] | 9 (42.9) | 22 (25.9) | 0.13 |
| Metabolic, genetic, or endocrine[i] | 4 (19.0) | 11 (12.9) | 0.49 |
| **Nirsevimab** | | | |
| First season nirsevimab | 14 (66.7) | 41 (48.2) | 0.13 |
| **Coinfections[j]** | | | |
| Respiratory viruses (n=106) | 6/21 (28.6) | 17/85 (20.0) | 0.39 |
| Influenza (n=95) | 2/17 (11.8) | 1/78 (1.3) | 0.08 |
| Human metapneumovirus (n=64) | 1/12 (8.3) | 2/52 (3.8) | 0.47 |
| Adenovirus (n=62) | 0/12 | 3/50 (6.0) | 0.61 |
| Parainfluenza (n=64) | 0/12 | 3/52 (5.8) | 0.62 |
| Rhinovirus/Enterovirus (n=64) | 3/12 (25.0) | 9/52 (17.3) | 0.68 |
| Bacterial pathogen (n=37)[j] | 3/9 (33.3) | 3/28 (10.7) | 0.28 |

Abbreviations: IQR, interquartile range; RSV, respiratory syncytial virus

a. Chi-squared p-value to assess differences in proportions across groups; comparisons with expected cell counts <5 in >20% of cells used Fisher's exact test
b. 2024-25 season among children admitted between November 6, 2024-April 7, 2025; 2025-26 season among children admitted October 2, 2025-March 31, 2026
c. Gestational age at birth was assessed among children <12 months of age at hospitalization
d. Eligibility criteria as of August 2026

e. Summary group of any of respiratory, cardiac, neurological/neuromuscular, and metabolic/genetic/endocrine conditions
f. Neurologic/Neuromuscular conditions included Muscular dystrophy, static encephalopathy, spastic quadriplegia, seizure disorders, neuromuscular weakness, and hydrocephalus
g. Respiratory conditions included asthma, requiring assistance with clearance of secretions, and upper airway disorders
h. Cardiac conditions included congenital heart disease, history of cardiac repair, pulmonary hypertension, and congestive heart failure
i. Metabolic, genetic, or endocrine conditions included trisomy 21, other chromosomal disorders, mitochondrial disorders, diabetes, and other metabolic disorders
j. Not all patients were tested for other pathogens; coinfection results are among those who received clinical testing for the specific pathogen; each row represents the number of patients tested for that specific pathogen
k. Represents culture results from blood (n=24), urine (n=8), and endotracheal (n=7) specimens collected from 40 patients.

Supplemental Table 2. Characteristics of case-patients with respiratory syncytial virus (RSV) and control-patients testing negative for RSV who did not have a codetection with influenza or SARS-CoV-2, and among those who did and did not receive second season nirsevimab, among nirsevimab eligible children between 8 and 23 months of age during their second RSV season, November 1, 2024-April 15, 2025 and September 15, 2025-April 15, 2026, Overcoming RSV Network

| Characteristic | Total No. (%) | RSV+ case patients No. (%) | RSV- control patients No. (%) | P-value[a] | Received nirsevimab No. (%) | Did not receive nirsevimab No. (%) | P-value[a] |
|---|---|---|---|---|---|---|---|
| **Total** | **211** | **103** | **108** | | **66** | **145** | |
| **RSV season[b]** | | | | | | | |
| 2024-25 season | 112 (53.1) | 58 (56.3) | 54 (50.0) | 0.36 | 36 (54.5) | 76 (52.4) | 0.77 |
| 2025-26 season | 99 (46.9) | 45 (43.7) | 54 (50.0) | | 30 (45.5) | 69 (47.6) | |
| **Age - median (IQR)** | 15.7 (13.4, 18.5) | 15.7 (14.0, 18.7) | 15.8 (13.0, 18.4) | 0.92 | 15.5 (12.7, 18.4) | 15.9 (13.8, 18.7) | 0.08 |
| **Sex** | | | | | | | |
| Male | 115 (54.5) | 59 (57.3) | 56 (51.9) | 0.43 | 36 (54.5) | 79 (54.5) | 0.99 |
| Female | 96 (45.5) | 44 (42.7) | 52 (48.1) | | 30 (45.5) | 66 (45.5) | |
| **Race and ethnicity[c]** | | | | | | | |
| Non-Hispanic White | 69 (32.7) | 30 (29.1) | 39 (36.1) | 0.64 | 25 (37.9) | 44 (30.3) | 0.30 |
| Non-Hispanic Black | 45 (21.3) | 24 (23.3) | 21 (19.4) | | 18 (27.3) | 27 (18.6) | |
| Non-Hispanic Asian | 8 (3.8) | 4 (3.9) | 4 (3.7) | | 2 (3.0) | 6 (4.1) | |
| Hispanic | 43 (20.4) | 25 (24.3) | 18 (16.7) | | 11 (16.7) | 32 (22.1) | |
| Multiple or other | 9 (4.3) | 3 (2.9) | 6 (5.6) | | 2 (3.0) | 7 (4.8) | |
| American Indian/Alaska Native | 23 (10.9) | 10 (9.7) | 13 (12.0) | | 3 (4.5) | 20 (13.8) | |
| Unknown | 14 (6.6) | 7 (6.8) | 7 (6.5) | | 5 (7.6) | 9 (6.2) | |
| **Region[d]** | | | | | | | |
| Northeast | 16 (7.6) | 8 (7.8) | 8 (7.4) | 0.13 | 4 (6.1) | 12 (8.3) | 0.38 |
| Midwest | 21 (10.0) | 15 (14.6) | 6 (5.6) | | 6 (9.1) | 15 (10.3) | |
| South | 106 (50.2) | 46 (44.7) | 60 (55.6) | | 39 (59.1) | 67 (46.2) | |
| West | 68 (32.2) | 34 (33.0) | 34 (31.5) | | 17 (25.8) | 51 (35.2) | |
| **Social vulnerability index – median (interquartile range)** | 0.5 (0.3, 0.7) | 0.5 (0.3, 0.7) | 0.5 (0.4, 0.7) | 0.59 | 0.5 (0.4, 0.7) | 0.5 (0.3, 0.7) | 0.32 |
| **Gestational age (n=28)[e]** | | | | | | | |
| <37 weeks gestation | 8/26 (30.8) | 3/11 (27.3) | 5/15 (33.3) | 0.74 | 3 (27.3) | 5 (33.3) | 0.74 |
| ≥ 37 weeks gestation | 18/26 (69.2) | 8/11 (72.7) | 10/15 (66.7) | | 8 (72.7) | 10 (66.7) | |
| **Nirsevimab eligibility[f]** | | | | | | | |

| | | | | | | | |
|---|---|---|---|---|---|---|---|
| Chronic lung disease of prematurity | 165 (78.2) | 86 (83.5) | 79 (73.1) | 0.07 | 61 (92.4) | 104 (71.7) | <.001 |
| Severe immunocompromise | 25 (11.8) | 7 (6.8) | 18 (16.7) | 0.03 | 3 (4.5) | 22 (15.2) | 0.03 |
| American Indian/Alaska Native | 23 (10.9) | 10 (9.7) | 13 (12.0) | 0.59 | 3 (4.5) | 20 (13.8) | 0.05 |
| Cystic fibrosis | 5 (2.4) | 3 (2.9) | 2 (1.9) | 0.68 | 1 (1.5) | 4 (2.8) | 0.58 |
| **Preexisting health conditions** | | | | | | | |
| Any underlying condition[g] | 132 (62.6) | 65 (63.1) | 67 (62.0) | 0.87 | 53 (80.3) | 79 (54.5) | <.001 |
| Neuromuscular or neurologic condition[h] | 45 (21.3) | 15 (14.6) | 30 (27.8) | 0.02 | 17 (25.8) | 28 (19.3) | 0.29 |
| Respiratory condition[i] | 85 (40.3) | 35 (34.0) | 50 (46.3) | 0.07 | 39 (59.1) | 46 (31.7) | <.001 |
| Cardiac condition[j] | 69 (32.7) | 29 (28.2) | 40 (37.0) | 0.17 | 32 (48.5) | 37 (25.5) | <.001 |
| Metabolic, genetic, or endocrine[k] | 35 (16.6) | 14 (13.6) | 21 (19.4) | 0.25 | 13 (19.7) | 22 (15.2) | 0.41 |
| **Nirsevimab** | | | | | | | |
| Nirsevimab in second season | 66 (31.3) | 19 (18.4) | 47 (43.5) | <.001 | 66 (100.0) | N/A | N/A |
| Days from nirsevimab receipt to symptom onset | 69.0 (49.0, 99.0) | 80.0 (64.0, 122.0) | 66.0 (31.0, 96.0) | <.001 | 69.0 (49.0, 99.0) | N/A | N/A |
| First season nirsevimab | 111 (52.6) | 53 (51.5) | 58 (53.7) | 0.74 | 37 (56.1) | 74 (51.0) | 0.50 |
| **Highest level of respiratory support** | | | | | | | |
| Low flow or no respiratory support | 90 (44.6) | 40 (40.0) | 50 (49.0) | 0.02 | 30 (50.0) | 60 (42.3) | 0.002 |
| High flow nasal cannula | 46 (22.8) | 24 (24.0) | 22 (21.6) | | 9 (15.0) | 37 (26.1) | |
| Noninvasive ventilation[l] | 39 (19.3) | 27 (27.0) | 12 (11.8) | | 6 (10.0) | 33 (23.2) | |
| Invasive mechanical ventilation | 27 (13.4) | 9 (9.0) | 18 (17.6) | | 15 (25.0) | 12 (8.5) | |
| **Clinical course** | | | | | | | |
| Death during hospitalization | 1 (0.5) | 0 | 1 (0.9) | 1.00 | 0 | 1 (0.7) | 1.00 |
| ICU admission | 98 (46.4) | 47 (45.6) | 51 (47.2) | 0.82 | 30 (45.5) | 68 (46.9) | 0.85 |
| ECMO | 1 (0.5) | 1 (1.0) | 0 | 1.00 | 1 (1.5) | 0 | |
| Hospital length of stay | 5.0 (4.0, 9.0) | 6.0 (5.0, 10.0) | 4.0 (3.0, 7.0) | <.001 | 5.0 (3.0, 9.0) | 5.0 (4.0, 8.5) | 0.92 |

Abbreviations: ECMO, extracorporeal membrane oxygenation; ICU, intensive care unit; IQR, interquartile range; RSV, respiratory syncytial virus

a. Chi-squared p-value to assess differences in proportions across groups; comparisons with expected cell counts <5 in >20% of cells used Fisher's exact test; comparisons of continuous variables used Kruskal-Wallis tests
b. 2024-25 season among children admitted between November 6, 2024-April 7, 2025; 2025-26 season among children admitted October 2, 2025-March 31, 2026
c. Statistical testing did not include American Indian/Alaska Native children
d. Northeast: Connecticut Children's Medical Center, Boston Children's Hospital, Columbia University, Children's Hospital of Philadelphia; Midwest: Ann & Robert H. Lurie Children's Hospital of Chicago, Riley Hospital for Children, Mayo Clinic Children's, University of Minnesota Masonic Children's Hospital, Children's Mercy Kansas City, Cincinnati Children's Hospital Medical Center; South: Children's of Alabama, Arkansas Children's Hospital, Nemours Children's Hospital, Children's Healthcare of Atlanta, Manning Family Children's, UNC Children's Hospital, Medical University of South Carolina, Monroe Carell Jr. Children's Hospital at Vanderbilt; West: Arizona Mesa Children's Hospital, UCSF Benioff Children's Hospital, Children's Hospital Colorado, Doernbecher Children's Hospital, Primary Children's Hospital, Rady Children's Hospital, Seattle Children's Hospital
e. Gestational age at birth was assessed among children <12 months of age at hospitalization
f. Eligibility criteria as of August 2026
g. Summary group of any of respiratory, cardiac, neurological/neuromuscular, and metabolic/genetic/endocrine conditions
h. Neuromuscular or neurologic conditions included muscular dystrophy, static encephalopathy, spastic quadriplegia, seizure disorders, neuromuscular weakness, and hydrocephalus
i. Respiratory conditions included asthma, requiring assistance with clearance of secretions, and upper airway disorders
j. Cardiac conditions included congenital heart disease, history of cardiac repair, pulmonary hypertension, and congestive heart failure
k. Metabolic, genetic, or endocrine conditions included trisomy 21, other chromosomal disorders, mitochondrial disorders, diabetes, and other metabolic disorders
l. Noninvasive ventilation defined as receipt of bilevel positive airway pressure or continuous positive airway pressure

Supplemental Figure 2. Second season nirsevimab product effectiveness against respiratory syncytial virus-associated hospitalizations among nirsevimab-eligible children aged 8-23 months during their second RSV season without codetections with influenza or SARS-CoV-2, November 1, 2024-April 15, 2025 and September 15, 2025-April 15, 2026, Overcoming RSV Network

| | Days from nirsevimab dose to symptom onset, Median (IQR) | No. Immunized Children / Total no. (%) | | Unadjusted Effectiveness % (95% CI) | Adjusted Effectiveness % (95% CI) |
|---|---|---|---|---|---|
| | | RSV+ case-patients | RSV- control patients | | |
| Overall effectiveness of nirsevimab[a] | 69.0 (49.0, 99.0) | 19/103 (18.4) | 47/108 (43.5) | 70.6 (45.1, 84.3) | 80.7 (59.2, 90.9) |
| Nirsevimab 8-59 days before symptom onset | 29.0 (20.0, 50.0) | 4/88 (4.5) | 19/80 (23.8) | 83.2 (49.7, 94.4) | 88.3 (59.2, 96.6) |
| Nirsevimab 60 or more days before symptom onset | 88.0 (70.0, 122.0) | 15/99 (15.2) | 28/89 (31.5) | 60.4 (19.7, 80.5) | 76.4 (45.7, 80.7) |
| Matched analysis[b] | | | | | |
| Any nirsevimab receipt | 69.0 (48.5, 97.5) | 13/64 (20.3) | 43/98 (43.9) | 61.7 (23.8, 80.8) | 71.5 (30.7, 88.3) |

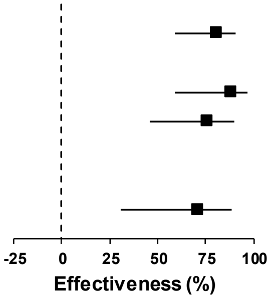

Abbreviations: CI, confidence interval; IQR, interquartile range; RSV, respiratory syncytial virus

a. Product effectiveness (PE, %) was calculated as (100*(1-adjusted odds ratio), with adjusted odds ratio calculated using unconditional logistic regression and adjusted for age, medical conditions other than nirsevimab, month/year of hospitalizations, and sex; controlled for site as a random effect. Data from the Overcoming RSV network are a case control study with a test negative design.

b. Product effectiveness (PE, %) was calculated as (100*(1-adjusted odds ratio), with adjusted odds ratio calculated using conditional logistic regression and adjusted for age, medical conditions other than nirsevimab, month/year of hospitalization, and sex. Data from the Overcoming RSV network are a case control study with a test negative design.
